# The ongoing COVID-19 epidemic in Minas Gerais, Brazil: insights from epidemiological data and SARS-CoV-2 whole genome sequencing

**DOI:** 10.1101/2020.05.05.20091611

**Authors:** Joilson Xavier, Marta Giovanetti, Talita Adelino, Vagner Fonseca, Alana Vitor Barbosa da Costa, Adriana Aparecida Ribeiro, Katlin Nascimento Felicio, Clara Guerra Duarte, Marcos Vinicius Ferreira Silva, Álvaro Salgado, Mauricio Teixeira Lima, Ronaldo de Jesus, Allison Fabri, Cristiane Franco Soares Zoboli, Thales Gutemberg Souza Santos, Felipe Iani, Ana Maria Bispo de Filippis, Marilda Agudo Mendonça Teixeira de Siqueira, André Luiz de Abreu, Vasco de Azevedo, Dario Brock Ramalho, Carlos F. Campelo de Albuquerque, Tulio de Oliveira, Edward C. Holmes, José Lourenço, Luiz Carlos Junior Alcantara, Marluce Aparecida Assunção Oliveira

## Abstract

The recent emergence of a previously unknown coronavirus (SARS-CoV-2), first confirmed in the city of Wuhan in China in December 2019, has caused serious public health and economic issues due to its rapid dissemination worldwide. Although 61,888 confirmed cases had been reported in Brazil by 28 April 2020, little was known about the SARS-CoV-2 epidemic in the country. To better understand the recent epidemic in the second most populous state in southeast Brazil (Minas Gerais, MG), we looked at existing epidemiological data from 3 states and sequenced 40 complete genomes from MG cases using Nanopore. We found evidence of multiple independent introductions from outside MG, both from genome analyses and the overly dispersed distribution of reported cases and deaths. Epidemiological estimates of the reproductive number using different data sources and theoretical assumptions all suggest a reduction in transmission potential since the first reported case, but potential for sustained transmission in the near future. The estimated date of introduction in Brazil was consistent with epidemiological data from the first case of a returning-traveler from Lombardy, Italy. These findings highlight the unique reality of MG’s epidemic and reinforce the need for real-time and continued genomic surveillance strategies as a way of understanding and therefore preparing against the epidemic spread of emerging viral pathogens.

## Introduction

The World Health Organization (WHO) office in China was informed about a cluster of new cases of pneumonia of unknown etiology in the City of Wuhan (Hubei province), in late December 2019 [1]. Shortly afterwards, a new type of coronavirus, now termed SARS-CoV-2, was isolated and identified by Chinese authorities, with its genetic sequence shared with the international community on 10 January 2020 [2–5]. Phylogenetic analysis revealed that SARS-CoV-2 was similar to other (pandemic) betacoronaviruses, such as severe acute respiratory syndrome coronavirus (SARS-CoV) and Middle East respiratory syndrome coronavirus (MERS-CoV) [4,5]; revealing also its phylogenetic relationship to other coronaviruses isolated from bats and Malayan pangolins (*Manis javanica*), indicating a likely zoonotic origin [2,5–7].

To date, more than 3.5 million cases of the disease caused by SARS-CoV-2, termed COVID-19, have been reported around the world [8,9]. On 11 March 2020, the WHO declared the COVID-19 a pandemic, prompting a dramatic increase in international concern and response [10]. On 26 February 2020, the first confirmed case of COVID-19 was reported in São Paulo (SP) state, Brazil [11]. Two months later (28 April 2020), 61,888 cases and 4,205 deaths attributed to COVID-19 had been reported in Brazil [12]. Meanwhile, preliminary phylogenetic analysis using the first two SARS-CoV-2 complete genomes isolated in São Paulo from travelers returning from Italy, revealed two independent introductions into the country, relative to the analyzed dataset available at that time [13].

The state of Minas Gerais (MG) is the second largest Brazilian state in terms of population size, estimated at approximately 21 million people, and is located near the state of São Paulo [14]. Due to its large population size and its well-connected and active neighboring states such as São Paulo and Rio de Janeiro, the state of MG is likely to be highly affected by the COVID-19 pandemic.

Genetic analyses and surveillance allow the characterization of circulating viral lineages, the inference of introduction events and the reconstruction of transmission patterns [15]. Together with epidemiological data, they are powerful tools to assist public health initiatives and preparedness. In this study, we present a summary of epidemiological data and the generation and analysis of 40 new SARS-CoV-2 genome sequences isolated from clinical samples of confirmed cases from MG, with the aim of providing a preliminary epidemiological overview of the circulation and introduction events of the virus in that state.

## Results/Discussion

After the WHO declared the outbreak of SARS-CoV-2 a Public Health Emergency of International Concern (PHEIC) on 30 January 2020, the Brazilian government declared a Public Health Emergency of National Importance on 3 February 2020, enabling the introduction of measures to prevent and control spread [16]. Twenty-three days later, the first confirmed case in Brazil was reported in the city of São Paulo, related to a traveler returning from Lombardy, Italy (Fig 1) [11]. By the 28 April 2020, more than 61,888 COVID-19 cases were confirmed in Brazil, 1,578 of which were from MG (Fig 2A) [17]. Over this period, MG registered 71 COVID-19-related deaths, and the capital city Belo Horizonte, with an estimated population of 2.5 million people, had reported 555 cases [17,18]. Fig 2A shows MG’s epidemic (reported cases) curve compared to the curves of two other neighboring states, São Paulo (SP) and Rio de Janeiro (RJ).

**Figure 1.**
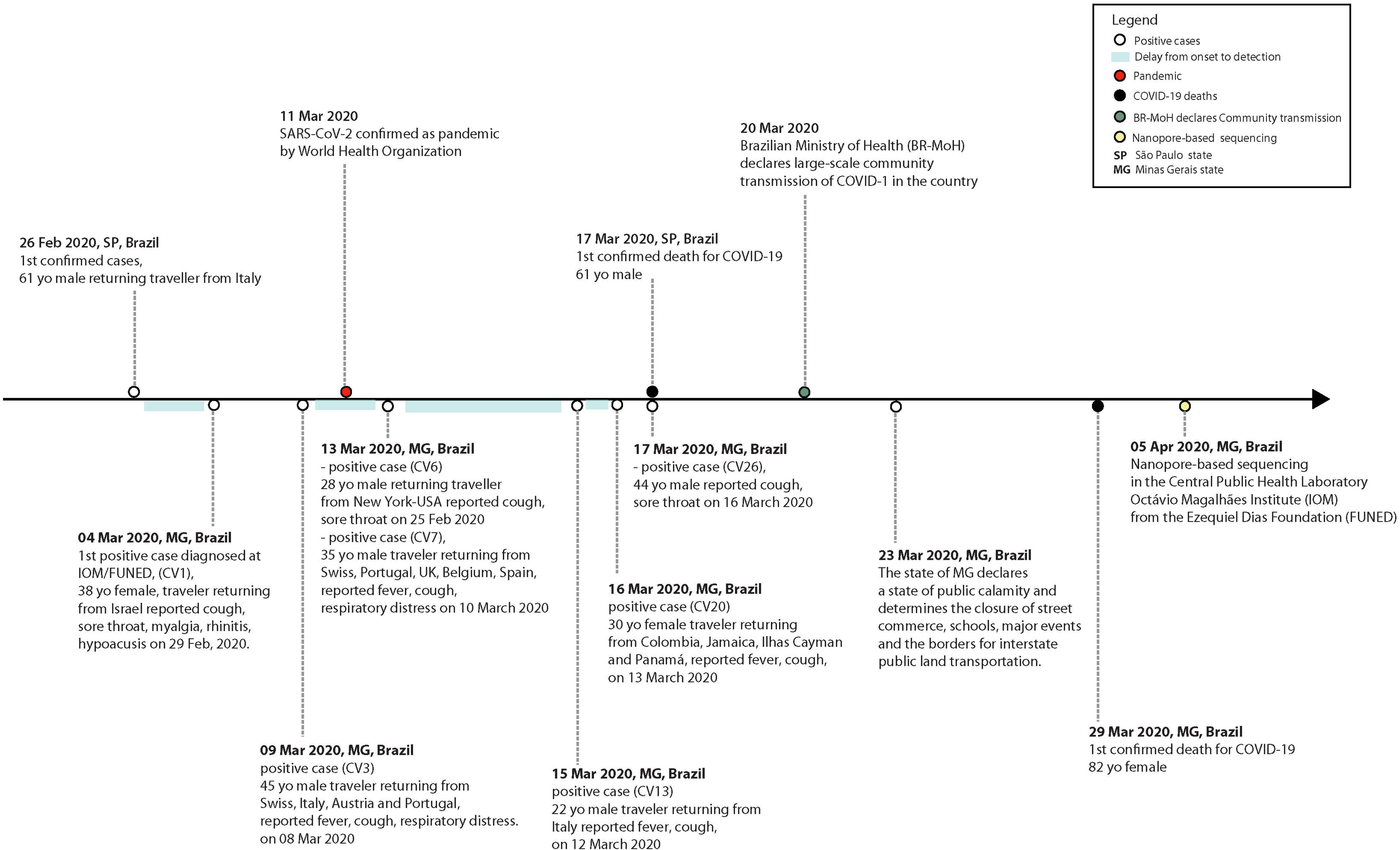
Timeline of key events following the confirmation of the first confirmed case of COVID-19 in Brazil. Events below the line occurred in Minas Gerais (MG) state, while national events are presented above the line. Codes in parentheses refer to the identification code (CV#) of the isolates from cases described in this study.

**Figure 2.**
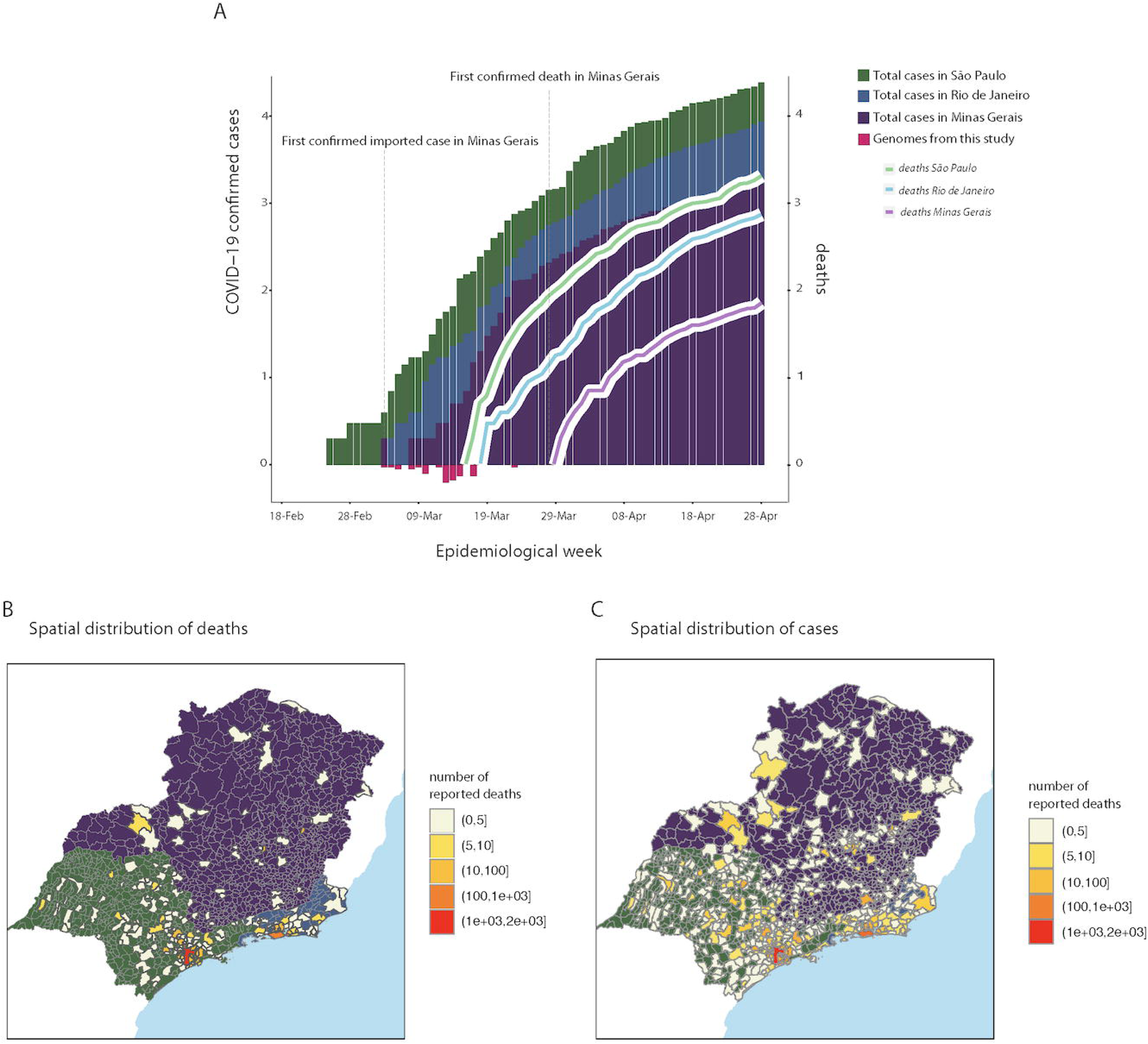
SARS-CoV-2 epidemic curve and spatial distribution of cases and deaths reported in the states of Minas Gerais (MG), São Paulo (SP) and Rio de Janeiro (RJ) Panel A: Daily confirmed cases of COVID-19 in the state of MG. The X axis represents the days from the first case in Brazil until 28 April 2020, while the Y axis represents the number of cases. The opposite side of the Y axis represents the number of deaths related to COVID-19. Numbers from Y axis are represented as log10. Panel B and C: Map with location (municipality) of deaths and case events, colored by total number of reports. Different background colors highlight the boundaries of the three states: Green for SP, purple for MG, Blue for RJ.

Without access to the total number of tests in time and in each state, we calculated the case fatality ratio (CFR) for MG, SP and RJ as the crude ratio between reported deaths and cases [19]. The CFR was found to increase with time in all states (S2 Fig), with means from date of first reported case up to the 28 April in each state at 2.67% for MG, 5.39% for RJ and 6.0% for SP. For SP and RJ, the CFR was consistently higher than reported elsewhere; for example at 2.6% (95% CI 0.89-6.7) for the Diamond Princess cruise ship [20], and 3.67% (95% CI 3.56-3.80) and 1.2% (95% CI 0.3-2.7) and 1.4% (95% CI 0.9-2.1) for Chinese regions [20–22].

We used the mortality time series (MTS) from MG, SP and RJ to project the cumulative number of infections, making two main simplifying assumptions: first, that the infection fatality ratio (IFR) of SARS-CoV-2 would be similar in the Brazilian states to that reported elsewhere; and second, that the number of cumulative deaths in each state were well reported. We considered the IFR estimated by Verity and colleagues (0.66%, CI 95% 0.39-1.33% [21]), for its general use in the modelling literature [23]. The cumulative number of infections in time is taken to be 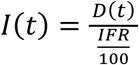, where *D(t)* is the cumulative number of deaths. From *I(t)* we further obtain the observation rate θ of reported cases from 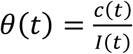 where *c(t)* is the number of reported cases in time. We found the observation rate to have decreased in time for all states - a likely outcome of successful tracing and testing only in the beginning, but with epidemic growth superseding tracing and testing efforts as the epidemic progressed (S1 Fig). By 28 April 2020, the last time point analyzed, RJ and SP had similar observation rates at 7.6% and 7.74% (respectively), while MG, where the epidemic started later, the observation rate was 15.3% (1 case in 7 infections).

To compare transmission potential, we used reported cases (CTS) and mortality time series (MTS) from MG, SP and RJ states to estimate the (effective) reproduction number R. For this, we performed maximum likelihood estimation of the (CTS and MTS) epidemic growth *r* using a phenomenological model, and two theoretical formulations on how *r* relates to R - one based on the SEIR epidemiological framework by Wallinga and colleagues [24], and another on the distribution of the serial interval [23] (see Supplementary Material for details). R was found to decrease in time since first reported case for all states (S5 and S10 Figs). When considering the entire period from first reported case to the 28 April, estimation methods gave similar R results per state (S6 and S11 Figs). For example, when using the CTS and serial interval formulation, R was 1.91 (CI 95% 1.2-3.1) for SP, 1.88 (CI 95% 1.27-2.8) for RJ and 1.82 (CI 95% 1.2-3.25) for MG.

When using geographic information from reported cases in each state (Fig 2C and S13-14 Figs), we found that cases were generally very dispersed in MG and more centralized around capital cities in RJ and SP. In MG, reported cases were on average ~103 km away (CI 95% 1.39-488) from the capital Belo Horizonte, while in SP they were ~0.05 km away from the city of São Paulo (CI 95% 0.05-269), and in RJ ~1.45 km away from the city of Rio de Janeiro (CI 95% 1.4-130). Similar patterns were found for reported deaths (Fig 2B and S15-16 Figs). In MG, reported deaths were on average ~229 km (CI 95% 1.39-488) away from Belo Horizonte, while in SP they were ~28 km from São Paulo (CI 95% 0.05-270), and in RJ ~18 km away from Rio de Janeiro (CI 95% 1.45-142).

Typically, incidence (cases, deaths) would be normalized per 100K individuals, taking into account the total population size of each state. Because of the very different spatial dispersion of cases and deaths in MG when compared to SP and RJ, we decided to also calculate the effective population size - the sum of the population sizes of all municipalities with reports. When using reported cases, we found that the effective population sizes were ~100%, ~100% and 64% of the total population sizes of RJ, SP and MG, respectively. When using reported deaths, the effective population sizes were ~95%, ~92%, and 35% of the total population sizes of RJ, SP and MG, respectively. Overall these numbers suggest that in MG cases and deaths have been reported only in a subset of the overall population, while in the other states SARS-CoV-2 appears widely dispersed. Incidence of reported cases per 100K using the effective population size was ~60 in SP, ~51 in RJ and ~7.85 in MG (S7 Fig), while incidence of deaths per 100K was ~5.56 in SP, ~4.69 in RJ and ~0.94 in MG (S12 Fig).

In MG, samples from (clinically) suspected cases were screened at the Central Public Health Laboratory/Octávio Magalhães Institute (IOM) of the Ezequiel Dias Foundation (FUNED), which belongs to the public laboratories network of the Brazilian Ministry of Health (MoH). As of 3 April 2020, IOM/FUNED had performed 3,303 RT-qPCR tests for SARS-CoV-2 on swab samples from suspected cases. We used Nanopore sequencing to generate complete genomes from 40 COVID-19 patients living in 15 different MG’s municipalities (Table 1).

**Table 1.** Information on the 40 sequenced samples from Minas Gerais state.

| Project-ID | Lab ID | Sample type | Ct value | Onset date | Collection date | Age | Sex | State | Municipality | Travel information |
| --- | --- | --- | --- | --- | --- | --- | --- | --- | --- | --- |
| CV1 | 47/20 | SWAB | 20.54 | 29/02/20 | 04/03/20 | 38 | F | MG | Ipatinga | Israel |
| CV2 | 115/20 | SWAB | 24.41 | 06/03/20 | 08/03/20 | 44 | F | MG | Sete Lagoas | Portugal, Spain |
| CV3 | 135/20 | SWAB | 27.77 | 08/03/20 | 09/03/20 | 45 | M | MG | Belo Horizonte | Italy, Switzerland, Austria, Portugal |
| CV4 | 242/20 | SWAB | 21.92 | N/A | 09/03/20 | 65 | M | MG | Juiz De Fora | USA |
| CV5 | 252/20 | SWAB | 29.93 | 12/03/20 | 12/03/20 | 32 | M | MG | Belo Horizonte | - |
| CV6 | 298/20 | SWAB | 18.69 | 13/03/20 | 13/03/20 | 28 | M | MG | Belo Horizonte | USA |
| CV7 | 352/20 | SWAB | 26.96 | 10/03/20 | 13/03/20 | 35 | M | MG | Belo Horizonte | Switzerland, Portugal, England, Belgium, Spain |
| CV8 | 399/20 | SWAB | 22.61 | 06/03/20 | 13/03/20 | 34 | M | MG | Belo Horizonte | - |
| CV9 | 428/20 | SWAB | 30.19 | 06/03/20 | 13/03/20 | 33 | F | MG | Belo Horizonte | Sao Paulo (Brazil) |
| CV11 | 607/20 | SWAB | 27.92 | 10/03/20 | 16/03/20 | 40 | F | MG | Mariana | Germany, Hungary, Czech Republic |
| CV12 | 615/20 | SWAB | 24.9 | 10/03/20 | 11/03/20 | 37 | F | MG | Juiz De Fora | USA |
| CV13 | 660/20 | SWAB | 25.69 | 15/03/20 | 15/03/20 | 22 | M | MG | Belo Horizonte | Italy |
| CV16 | 791/20 | SWAB | 20.64 | 15/03/20 | 16/03/20 | 52 | M | MG | Belo Horizonte | - |
| CV17 | 809/20 | SWAB | 26.95 | 09/03/20 | 11/03/20 | 61 | M | MG | Sete Lagoas | Portugal, Spain |
| CV18 | 833/20 | SWAB | 24.54 | 11/03/20 | 16/03/20 | 25 | F | MG | Belo Horizonte | - |
| CV19 | 836/20 | SWAB | 22.04 | 11/03/20 | 16/03/20 | 22 | M | MG | Belo Horizonte | Rio de Janeiro (Brazil) |
| CV20 | 838/20 | SWAB | 23.52 | 13/03/20 | 16/03/20 | 30 | F | MG | Belo Horizonte | Colombia, Jamaica, Cayman Islands, Panama |
| CV21 | 842/20 | SWAB | 16.67 | 05/03/20 | 16/03/20 | 56 | F | MG | Bom Despacho | - |
| CV22 | 895/20 | SWAB | 27.92 | 15/03/20 | 16/03/20 | 20 | F | MG | Mariana | Germany |
| CV24 | 1028/20 | SWAB | 25.15 | 13/03/20 | 16/03/20 | 22 | F | MG | Uberlândia | - |
| CV26 | 1078/20 | SWAB | 18.76 | 16/03/20 | 17/03/20 | 44 | M | MG | Belo Horizonte | - |
| CV27 | 1166/20 | SWAB | 22.99 | 11/03/20 | 17/03/20 | 60 | F | MG | Boa Esperança | - |
| CV28 | 1142/20 | SWAB | 22.21 | 13/03/20 | 17/03/20 | 46 | F | MG | São João Del Rei | USA |
| CV31 | 1274/20 | SWAB | 17.38 | 16/03/20 | 17/03/20 | 35 | M | MG | Betim | - |
| CV32 | 1290/20 | SWAB | 16.41 | 17/03/20 | 17/03/20 | 27 | M | MG | Betim | - |
| CV33 | 1420/20 | SWAB | 18.79 | 14/03/20 | 17/03/20 | 35 | M | MG | Sabara | - |
| CV34 | 1467/20 | SWAB | 22.31 | 16/03/20 | 18/03/20 | 48 | F | MG | Belo Horizonte | - |
| CV35 | 1500/20 | SWAB | 24.06 | 07/03/20 | 18/03/20 | 75 | M | MG | Poços De Caldas | Chile, Peru |
| CV36 | 1504/20 | SWAB | 24.07 | 18/03/20 | 18/03/20 | 50 | F | MG | Muriae | - |
| CV40 | 1834/20 | SWAB | 23.97 | 18/03/20 | 19/03/20 | 29 | M | MG | Belo Horizonte | - |
| CV41 | 1892/20 | SWAB | 22.84 | 16/03/20 | 18/03/20 | 20 | F | MG | Serra Do Salitre | - |
| CV42 | 2119/20 | SWAB | 18.78 | 18/03/20 | 20/03/20 | 67 | M | MG | São João Del Rei | - |
| CV43 | 2159/20 | SWAB | 24.81 | 14/03/20 | 17/03/20 | 19 | F | MG | Patrocínio | - |
| CV44 | 2196/20 | SWAB | 23.47 | 17/03/20 | 18/03/20 | 19 | F | MG | Patrocínio | - |
| CV45 | 2241/20 | SWAB | 22.85 | 14/03/20 | 20/03/20 | 58 | M | MG | Muriae | Sao Paulo (Brazil) |
| CV46 | 2271/20 | SWAB | 22.9 | 19/03/20 | 20/03/20 | 35 | M | MG | Belo Horizonte | - |
| CV47 | 2288/20 | SWAB | 22.4 | 17/03/20 | 19/03/20 | 35 | M | MG | Belo Horizonte | - |
| CV48 | 2693/20 | SWAB | 22.43 | 19/03/20 | 20/03/20 | 74 | M | MG | Varginha | - |
| CV49 | 2801/20 | SWAB | 20.95 | 16/03/20 | 22/03/20 | 30 | M | MG | Belo Horizonte | - |
| CV50 | 5068/20 | SWAB | 31.86 | 20/03/20 | 26/03/20 | 44 | M | MG | Mariana | - |

Project-ID=sample identifier; Onset date= Symptoms onset date; F=Female; M=Male; MG=State of
Minas Gerais; N/A=Not Available.

Of the 40 samples, 17 (42.50%) were from the state’s capital (Belo Horizonte), while the other municipalities were represented by one, or a maximum of three samples. These samples were from 17 females and 23 males, with a collection date ranging from 4 March 2020, from the first positive case diagnosed at IOM/FUNED, to 26 March 2020 (Table 1). The median age of the patients was 35 years (ranging from 19-79 years old). Selected samples had cycle threshold (Ct) values that ranged from 16.41 to 31.86 (median= 22.945). We found no demographic variables (age, gender) to be statistically correlated with sample Ct (S17 Fig). The new sequences have a median genome coverage of 82.5% related to the reference genome NC_045512.3 (S1 Table). All sequences generated in this study have been submitted to the GISAID Initiative following the WHO guidelines on the importance of sharing genomic data during situations of public health emergency of international concern [25].

Of the 17 (42.5%, n=40) sequenced cases with available travel history information, 14 cases (82.35%, n=17) reported international travel and three reported domestic travel. Two among the later visited the city of São Paulo and one the city of Rio de Janeiro (Table 1). Of the international travel-related cases, seven (50%) were linked to travel to European countries (Portugal, Spain, Italy, Switzerland, Austria, England, Belgium, Germany, Czech Republic, and Hungary), while six reported travel to countries in the Americas (USA, Colombia, Jamaica, Cayman Islands, Panama, Chile, and Peru). One case reported travel to Israel.

To explore the history of the virus in MG, we performed a maximum likelihood (ML) phylogenetic analysis on the dataset containing the 40 new sequences plus other 3,062 sequences deposited in GISAID up to 15 April 2020. Our estimated ML phylogeny identified two major clades branching at the root of the tree (Fig 3). These two clades were named lineages A and B, following a SARS-CoV-2 lineage nomenclature recently proposed [26].

**Figure 3.**
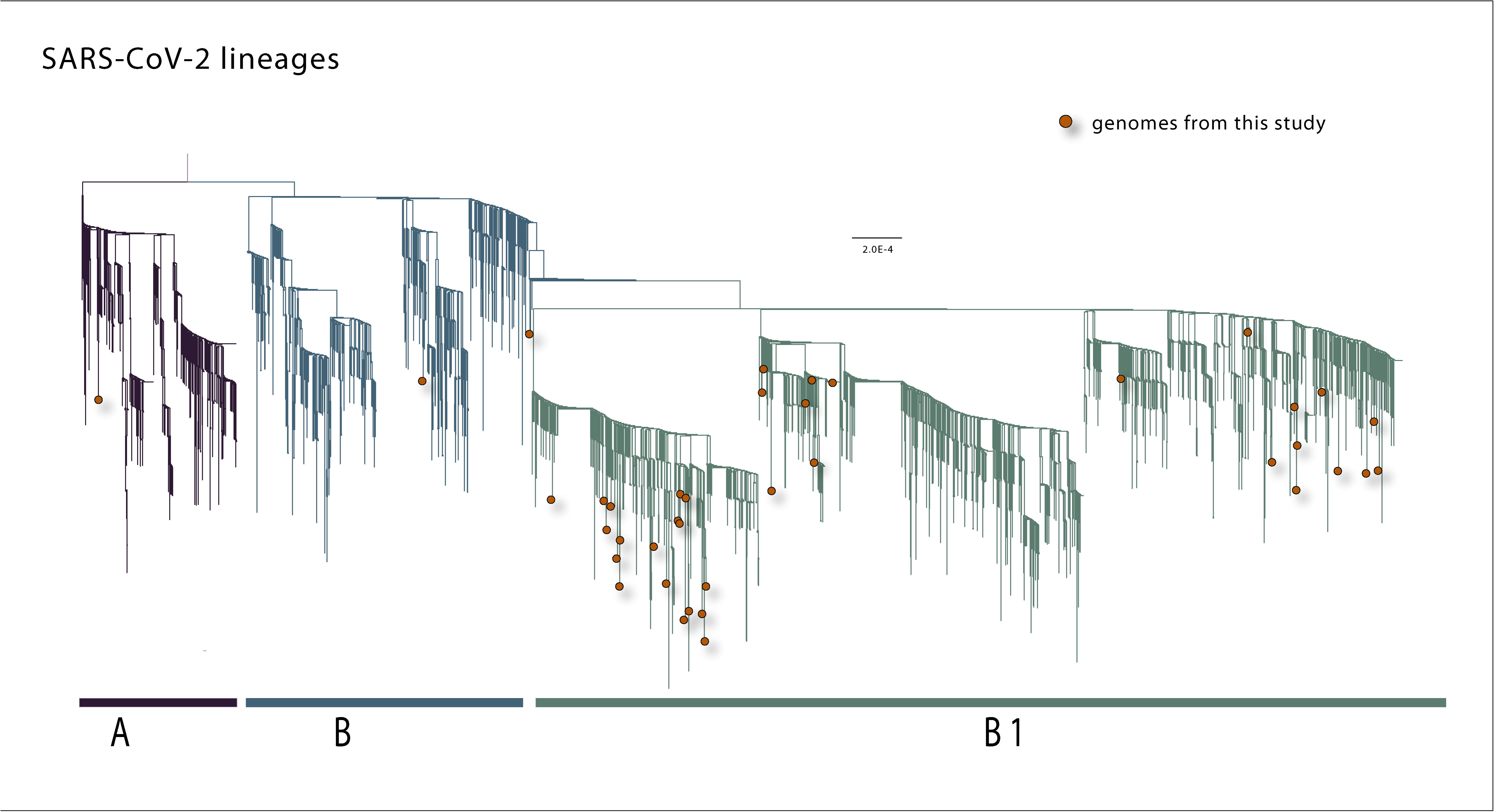
Phylogenetic analysis of the SARS-CoV-2 isolated in the state of MG, Brazil. A Maximum likelihood phylogeny inferred using 40 genome sequences from SARS-CoV-2 generated in this study and 3062 sequences already deposited in GISAID.

According to this nomenclature scheme, two main SARS-CoV-2 lineages could be identified as lineage A, defined by the Wuhan/WH04/2020 strain, and as lineage B represented by the Wuhan-Hu-1 strain. From these two main lineages, other sub-levels of descending lineages could be determined. Following the publication of this proposed lineage nomenclature scheme, a tool for automated lineage assignment was made publicly available in the GitHub repository (https://github.com/hCoV-2019/pangolin) [27]. We used this tool to perform the assignment of MG’s sequences to the lineages [26]. The results of this lineage assignment showed that the majority (n=32, 80%) of MG sequences were assigned to lineage B.1. This also includes sequences from other countries such as Australia, China, Canada, Malaysia, and USA [28]. Moreover, two sequences were assigned to lineage B.2 (isolates CV22 and CV36), and one sequence to lineage A (isolate CV7) (see S2 Table for full results).

Slightly different from the lineage assignment approach mentioned before, in our ML phylogeny most of MG’s new sequences (n=37, 92,5%) were placed in a descendant lineage we named B.1, which also included other sequences from GISAID sampled worldwide. Of these 37 sequences from MG within lineage B.1 (Fig 3), 11 are isolates from cases that reported travel to European countries (isolates CV2, CV3, CV11, CV13, CV17) or the Americas (isolates CV4, CV6, CV12, CV20, CV28, CV35), in addition to the isolate CV1 from a traveler who returned from Israel. Two MG’s sequences (CV22 and CV9) fell into lineage B, one of which (CV22) reported travel to Germany. The only sequence from MG that fell into lineage A refers to a case (CV7) that reported travel to European countries (Fig 3 and Table 1).

To assess these lineages in more detail and in time, we performed Bayesian time-measured phylogenetic analysis using a molecular clock model. We analyzed three sub-datasets (named subset A, subset B and subset B.1) extracted from each lineage from the ML tree that included Brazilian sequences. Our maximum clade credibility (MCC) trees showed that most of MG’s sequences were interspersed with other isolates sampled from other countries (Fig 4b, c, d). This pattern, similar to that observed in other countries [28–30], is also in accordance with our ML tree and with the epidemiological data, indicating that these isolates were linked to travel exposure rather than community transmission, and reinforcing the idea that multiple independent introductions with source abroad have occurred in MG.

**Figure 4.**
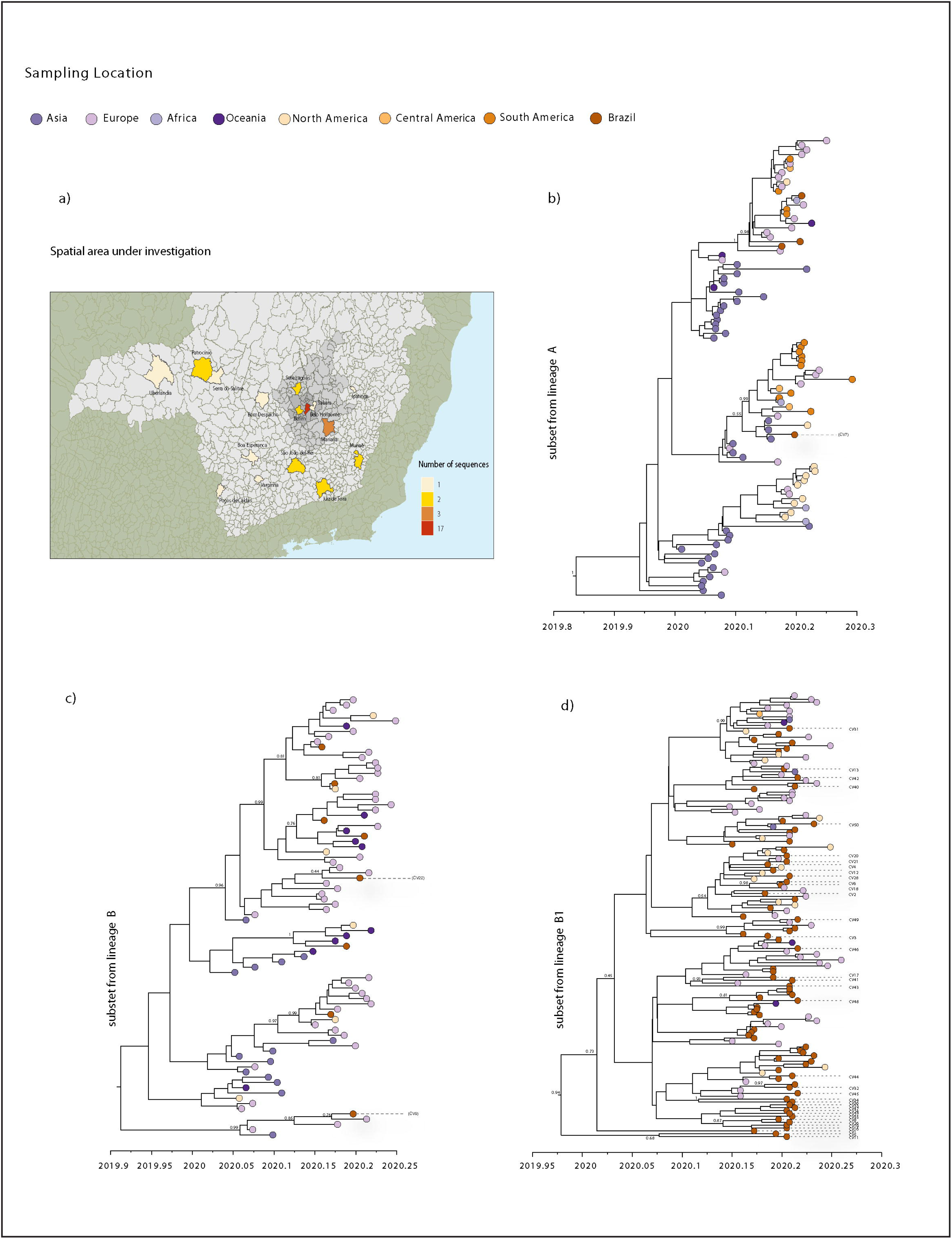
Bayesian analysis of the SARS-CoV-2 isolated in the state of MG, Brazil. (a) Map of the MG state showing the number of SARS-CoV-2 new sequences by patient’s municipality. b) Molecular clock phylogeny of the subset from lineage A, including one new sequence from MG. c) Molecular clock phylogeny of the subset from lineage B, including two new sequences from MG. d) Molecular clock phylogeny of the subset from lineage B.1, including 37 new sequences from MG. For molecular clock phylogenies, numbers along branches represent posterior probabilities and colors represent different sampling locations.

Despite the observed dispersed distribution, some sequences from MG grouped together, forming clusters that also included sequences from Brazil and other countries. Subset B.1 tree shows these clusters containing more than one sequence from MG (Fig 4d). However, these clusters have very low posterior probability support, because of the low genetic diversity of SARS-CoV-2 genomes currently available worldwide [31–33]. Nonetheless, four clusters, each consisting of only two MG sequences showed posterior probabilities of >80%. One of these clusters (Fig 4d), with a posterior probability of 100%, was formed by isolates CV34 and CV36, referring to cases of seemingly local transmission from contacts with a COVID-19 confirmed and suspected case, respectively.

From the time scaled phylogeny, we estimated the mean time of the most recent common ancestor (tMRCA) of the SARS-CoV-2 epidemic in Brazil to be 10 February 2020 (95% HPD interval 27 January to 22 February 2020), which is consistent with the start of reported cases in Brazil and with the epidemiological data from the first case confirmed in SP, regarding a traveler returning from Lombardy, Italy, on 21 February 2020 [11,13].

Despite the grouping of some MG sequences, we cannot infer a close relationship between these sequences with certainty at this stage, because of the small sample size data which covers only about 30 days of the epidemic in MG. That is, this dataset cannot fully represent the genetic diversity of SARS-CoV-2 strains circulating. Moreover, the low genetic diversity of sequences available so far limits conclusions about SARS-CoV-2 directionality and spread based solely on genetic data. As observed in another study [32], due to the described limitations of the available genomic data, the phylogenetic results presented should be approached with caution and considered as hypothesis-generating on the transmission events of SARS-CoV-2 in a local setting.

In conclusion, at the end of April 2020, the COVID-19 epidemic in the state of MG was still expanding (R>1) and it is highly dispersed with cases and deaths reported mostly away from the capital city and with approximately only 64% and 35% of the total population being represented in case and death reported data, respectively. Genomic data and other epidemiological information from travel-related cases, allowed us to identify several introduction events that occurred independently in MG, further helping to explain the geographical patchiness of reported cases and deaths. These initial insights based on the restricted data that is available show that transmission is likely to continue in the near future and suggest room to improve reporting. Increasing COVID-19 testing and SARS-CoV-2 genomic sequencing would help to better understand on how the virus is spreading and would thus inform better control of the COVID-19 epidemic in Brazil.

## Methods

### Ethics statement

Anonymized samples processed in this study were sent to the Central Public Health Laboratory/Octávio Magalhães Institute (IOM) of the Ezequiel Dias Foundation (FUNED), which belongs to public laboratories network from Brazilian Ministry of Health (BMoH). They were previously obtained by the local health services for routine diagnosis of SARS-CoV-2 and epidemiological surveillance. The availability of these samples for research purposes during outbreaks of international concern is allowed to the terms of the 510/2016 Resolution of the National Ethical Committee for Research - Brazilian Ministry of Health (CONEP - Comissão Nacional de Ética em Pesquisa, Ministério da Saúde), that authorize the use of clinical samples collected in the Brazilian Central Public Health Laboratories to accelerate knowledge building and contribute to surveillance and outbreak response.

### Sample collection and RT-qPCR diagnosis

Samples used in this study were residual anonymized clinical samples, with no or minimal risk to patients, provided for research and surveillance purposes as described above. Swab samples collected from COVID-19 suspected cases were sent from throughout the state of MG to IOM-FUNED facilities. At IOM-FUNED, these samples were submitted to total RNA extraction with an automated protocol on the QIAsymphony platform using QIAsymphony DSP Virus/Pathogen Kit (Qiagen), following the manufacturer’s recommendations. The molecular diagnosis was performed on a 7500 Real-Time PCR System (Thermofisher Scientific), using a RT-qPCR singleplex kit for the SARS-CoV-2 *envelope* and *RNA-dependent RNA polymerase* genes, developed by Bio-Manguinhos/Fiocruz (Rio de Janeiro, Brazil) and provided by the Brazilian Ministry of Healthy, following the manufacturer’s recommendations. We selected 48 samples with RT-qPCR positive results available until 3 April 2020 from patients residing in different municipalities of the state of MG and presenting symptoms such as fever, cough, headache, dyspnea, sore throat and/or vomiting.

Samples were selected based on the Ct value ≤ 32. Epidemiological data, such as symptoms, travel history and municipality of residency, was collected from medical records accompanying the collected samples provided by IOM/FUNED.

### cDNA synthesis and sequencing multiplex PCR

For the complementary DNA synthesis stage, the SuperScript IV Reverse Transcriptase kit (Invitrogen) was used following the manufacturer’s instructions. The generated cDNA generated was subjected to sequencing multiplex PCR using Q5 High Fidelity Hot-Start DNA Polymerase (New England Biolabs) and a set of specific primers, designed by ARTIC Network (https://github.com/artic-network/artic-ncov2019/tree/master/primer_schemes/nCoV-2019/V1) for sequencing the complete genome of SARS-CoV-2 [34]. PCR conditions have been previously reported in [34]. All experiments were performed on cabinet safety level 2.

### Whole genome sequencing

Amplified PCR products were purified using the 1x AMPure XP Beads (Beckman Coulter) following previously published protocol [35]. Purified PCR products were quantified using the Qubit® dsDNA HS Assay Kits (Invitrogen), following the manufacturer’s instructions. Of the 48 samples, only 40 contained enough DNA (≥ 2ng/μL) to proceed to library preparation. Sequencing libraries were prepared using the Oxford Nanopore Ligation Sequencing Kit (SQK-LSK109) following previously published protocol [35]. Before pooling all samples, each sample was barcoded using the Native Barcoding Expansion kits (NBD104 and EXP-NBD114). After barcoding adaptor ligation, sequencing libraries were loaded on a flow cell (FLO-MIN106) for subsequent MinION sequencing, programmed to run for six hours. Reads were basecalled using Guppy and barcode demultiplexing was performed using qcat. Consensus sequences were generated by *de novo* assembling using Genome Detective and Coronavirus Typing Tool [36,37].

### Phylogenetic analysis

Public SARS-CoV-2 complete genome sequences available up to 15 April 2020 were retrieved from the GISAID. Sequences were aligned using MAFFT (FF-NS-2 algorithm) following default parameters [38]. The alignment was manually curated to remove artifacts at the ends and within the alignment using Aliview [39]. Phylogenetic analysis of these genome sequences was performed using IQ-TREE (version 1.6.10) under the best fit model according to Bayesian Information Criterion (BIC) indicated by the Model Finder application implemented in IQ-TREE [40]. The statistical robustness of individual nodes was determined using 1000 bootstrap replicates.

Lineages assessment was conducted using Phylogenetic Assignment of Named Global Outbreak LINeages tool available at https://github.com/hCoV-2019/pangolin [27]. Four complete or near-complete SARS-CoV-2 genome datasets were generated. Dataset 1 (*n* =3,102) comprised the data reported in this study (*n* = 40) plus publicly available SARS-CoV-2 sequences (*n* = 3,062) from GISAID. Subsequently, to investigate the dynamic of the SARS-CoV-2 infection within the three different SARS-CoV-2 lineages (A, B and B.1), Bayesian molecular clock analysis was conducted on three smaller sub-datasets for each of the three lineages identified in the ML phylogeny and containing MG’ isolates (dataset 2 *for* subset A *n* = 100; dataset 3 *for* subset B *n* = 84; dataset 4 *for* subset B.1 *n* = 169). ML trees from these three sub-datasets were inspected in TempEst v1.5.3 for presence of temporal signal [41]. Linear regression of root-to-tip genetic distance against sampling date indicated that the SARS-CoV-2 sequences evolve in clock-like manner (r = 0.43; r = 0.47; r = 0.40 from subset A; B and B.1, respectively) (results in S18 Fig). For Bayesian time-scaled phylogenetic analysis we used BEAST 1.10.4 [42]. We employed the strict molecular clock model, which assumes a single rate across all phylogeny branches. We used the HKY+G4 codon partitioned (CP)1+2,3 substitution model and the exponential growth coalescent model [43]. We computed MCMC (Markov chain Monte Carlo) triplicate runs of 100 million states each, sampling every 10.000 steps for each sub-dataset. Convergence of MCMC chains was checked using Tracer v.1.7.1 [44]. Maximum clade credibility trees were summarized from the MCMC samples using TreeAnnotator after discarding 10% as burn-in.

## Data Availability

SARS-CoV-2 genome sequences generated in this study have been deposited in the GISAID platform (https://www.gisaid.org/), accession numbers IDs EPI_ISL_429664 to EPI_ISL_429703.

## Epidemiological data assembly

Data used in the epidemiological analysis were retrieved from https://github.com/wcota/covid19br [45].

## Acknowledgments

We would like to thank all the authors who have kindly deposited and shared genome data on GISAID. A table with genome sequence acknowledgments can be found in S3 Table. We thank all personnel from Health Surveillance System from the state of MG that helped with epidemiological data collection. We are also grateful for the support provided by the personnel from the Central Public Health Laboratory/Octávio Magalhães Institute (IOM) of the Ezequiel Dias Foundation (FUNED).

## Author contributions

**Conceptualization:** LCJA, MG, JL, JX and MAAO; **Data Curation:** JX; MG; VF; TdO; JL; TO; EH; and LCJA; **Formal Analysis:** JX; MG; VF; TdO; and JL; **Investigation:** JX; MG; VF; TA; AVBC; AAR; FI; KNF; CGD; MVFS; CFSZ; TGSS; MTL; AF; AF; AMBF; MAMTS; VA; LCJA and MAAO; **Validation:** JX; MG; JL; TdO, LCJA and MAAO; **Writing – Original Draft Preparation:** JX; MG; JL; TO; LCJA and MAAO; **Writing – Review & Editing:** JX; MG; TA; JL; TdO, LCJA and MAAO.

